# Reevaluation of the Variable Component of the Systematic Error Calls for Paradigm Change in Clinical Laboratory Quality Control

**DOI:** 10.1101/2023.05.24.23290382

**Authors:** Atilla Barna Vandra

## Abstract

The existence of the variable component of the systematic error (VCSE) is known since 1963, but seems to be a kind of taboo: neither has definition in VIM, nor is present in equations, being considered transformed in time into random error. Present study using methods of mathematical statistics, computer simulations and examples from the day-to-day practice of the author makes an attempt to reevaluate its role and significance in the QC in clinical laboratory.

“The bias” (which one?) it is a definitional uncertainty, because it is time-variable. Making clear distinction between bias measured in repeatability respective reproducibility within laboratory (RW) conditions as in case of standard deviations, and also separating constant and variable subcomponents of the systematic error, two sets of error parameters are obtained each set being consistent with the measurement conditions. The link between them is the time-variable VCSE function.

The conditions of calculations and predictions based on them must be consistent with the conditions of their determination, to avoid the redundant use. Being consequent, several differences can be discovered between the constant and the variable component of the systematic error respective random phenomena, also between variable and random error components.

The variable components of the systematic errors are cyclical, mostly predictable variations (shifts and drifts) in the daily mean. However, it is presented a direct method to determine the SD of the VCSE, its easiest calculation is possible from accurate values of s_r_ and s_RW_. There are presented methods to calculate s_r_ and s_RW_ from long term data.

As conclusion there is necessary a new paradigm of the QC in clinical laboratory based on the proposed error model.

**Motto:** *“Everything Should Be Made as Simple as Possible, But Not Simpler” (A. Einstein)*.

## Introduction

The existence variable component of the systematic error (VCSE) it is known but seems to be a kind of taboo, because has no definition in the VIM, it is not present in equations, and is considered incorrigible as RE ([1] R. Kadis, 2018). Despite it is the single error component which can be corrected by calibration, (although it cannot be definitively eliminated), it was hidden in the s_RW_, being considered transformed in time into random phenomena. ([2] E. Theodorsson & al. 2014). A real transformation does not happen, the VCSE is only measured together with real random error component, when calculating s_RW_. Such contradictions and the observation, that the VCSE frequently is included in s_RW_ without being eliminated from the bias motivated the reevaluation of its importance.

## Materials and methods

Present study it is a theoretical one, based on mathematical statistics methods. Most statements and observations are present in the literature, but only as mosaic pieces. Critical statements are based on theoretical deductions, computer simulations and observations made in 40-years experience of the author in clinical laboratory. Exemplifications are from day-to-day experience of the author in the laboratory of the Brasov County Clinical Hospital for Urgencies (SCJUBv), determinations were made on Cobas 6000 and Cobas pro analyzers with Roche reagents. No patient data was used.

### Theoretical background

There are two points of view in the clinical laboratory:

- The long-term point of view of the accreditation services common with that of the clinicians, who are interested mainly in the degree of the reliability of the measurement results, quantified as measurement uncertainty, the intervals in which the results can be found with a given confidence.
- A short-term (within day, intra-run) point of view of the laboratory specialist, who must take day-to-day (run-to run) decisions if in that day (run) he/she can run patient samples, or he/she must make supplementer maintenance, recalibrations or reagent changes before to avoid erroneous results of patient samples.

Each point of view is consistent with its area of applicability, neither of them is more correct in all situations. The error parameters (e. g. SD or bias) must be determined in the same conditions they will be used in calculations and predictions. E. g. an error parameter determined in short term conditions, leads to erroneous conclusions in long term decisions and vice versa.

The time-dependence of bias it is known since 1963 ([3] Ch. Eisenhart, 1963), however only in last years has got the deserved attention, and were made the first attempts to separate the bias of the moment, and the long term mean bias. ([4] A. B. Vandra, 2014)

In the present study, to be consistent with the usual notations of the measured standard deviations (s_r_ and s_RW_) a clear distinction was made between:

- Bias (B) measured in short term, (within day/run) constant, repeatability (‘r’) condition which will be noted B_r_(t) (highlighting its time-variable function character) called by W. P. Oosterhuis & al ([5] W. P. Oosterhuis & al. 2018) ‘within-day bias’, and
- Bias measured in long-term, variable, reproducibility within laboratory (RW) conditions noted B_RW_, called by W. P. Oosterhuis & al. ‘long term bias’ (without using different notation for them). B_RW_ is the long term mean of the daily B_r_(t) values.

‘Bias of the day’ ([4] A. Vandra, 2014) “short term bias’ ([5] W. P. Oosterhuis & al., 2018) and ‘bias measured in repeatability conditions’ (in present study), respective ‘mean bias’ ([4] A. Vandra, 2014), ‘long term bias’ ([5] W. P. Oosterhuis & al., 2018) and ‘bias measured in RW conditions’ (present study) are equivalent names for the same terms.

“The bias” it is a definitional uncertainty, because the time-variable character of the bias.

According to Ivo Leito:

“*Bias determined within a single day is different from one determined on different days (and averaged)”* ([6] I. Leito)

The time-variability of bias justifies the former distinction.

The definition of the systematic error component (SE) in VIM ([7], VIM, 2.17) is:

“The s*ystematic error component (SE) it is the component of measurement error that in replicate measurements remains constant* ***or*** *varies in a predictable manner”*.

The former definition by the ‘or’ word defines two subcomponents of the SE:

- The constant component of the SE (noted CCSE) which can be identified with the B_RW_ (the long-term mean of the daily biases),
- The variable component of the SE (VCSE(t) – also a time-variable function) which can be identified with the difference between the bias of the moment ‘t’ (measured in repeatability conditions) B_r_(t) and B_RW_ (the CCSE).

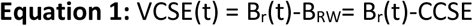

An analogue equation it is presented in Nordtest NT TR 537 ([8] B. Magnusson & al. p. 18), for the relationship between the SD-s measured in repeatability respective reproducibility within laboratory conditions (s_r_ respective s_RW_). The link between them it is also the VCSE(t), but expressed as a SD (s_VCSE_). A SD is not necessarily linked to the normal distribution. It can be calculated from any set of variable values, e. g. from random ones or from functions, so also from the VCSE(t) function.

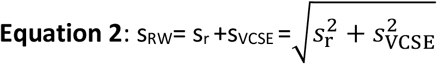

Regrouping the terms, can be calculated the s_VCSE_:

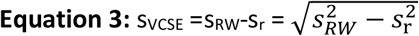

The authors of Nordtest NT TR 537 reduce the VCSE(t) to the „differences in calibration over time.” The gradual changes in daily means (drifts caused by the reagent property changes in time) also have an important contribution to the VCSE(t).

Although the CLSI EP15-A3 recommended method for estimation of precision and trueness (using 5×5 measurements) (CLSI, [9] followed by an ANOVA analysis of variance calculates the “between run variance” gives even the opportunity to calculate a short period s_VCSE_ as the square route of the between run variance, but these calculations did not become a practice, because it was neglected the importance of this error parameter. This opportunity it is another proof for its existence.

### Two points of view, two sets of error parameters

By making clear distinction between bias types, as we are accustomed in case of SD-s, two sets of error parameters are obtained. As was mentioned before, the conditions in which the error parameters are obtained must be consistent with the conditions, for which the calculations and predictions are made. A parameter measured in short term repeatability conditions cannot be used for long term calculations, in RW conditions, and vice-versa. E. g. EQA measurements are made in a single day, which means repeatability conditions, therefore only the bias of that day (B_r_(t)) can be obtained, B_RW_ not. Because the time-variability of the bias, the obtained value has only 24 hours term of validity, and cannot be used for long term decisions or calculations, e. g. to determine MU. On the other hand, based on long time-frame parameters determined in RW conditions cannot be taken accurate short-term decisions in internal quality control. This is a thought-provoking even shocking conclusion, because Westgard rules are based on s_RW_… The principles of the actual internal quality control must be reconsidered. Table 1 presents the error parameters, their determination conditions and the area of their applicability. All equations are well-known, what differs from their usual form with definitional uncertainties, it is which of the error components (measured in repeatability respective RW conditions) can be substituted in them.

**Table 1:**
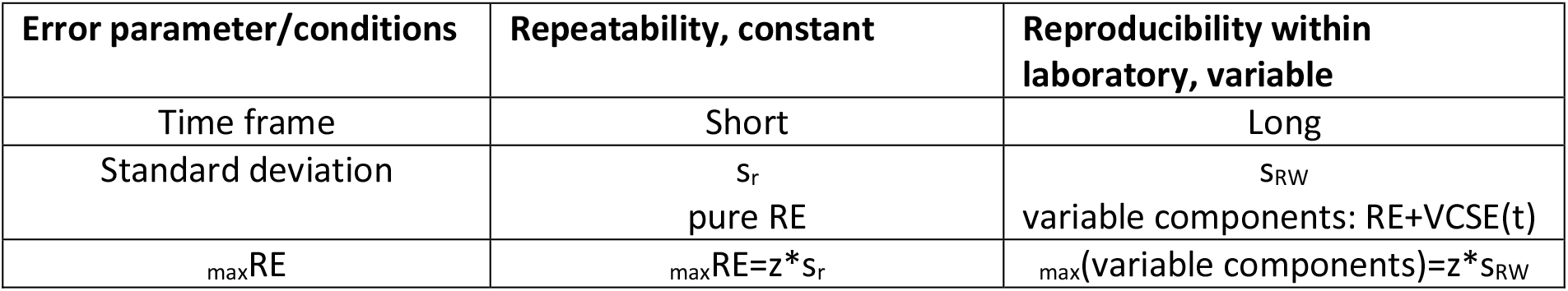
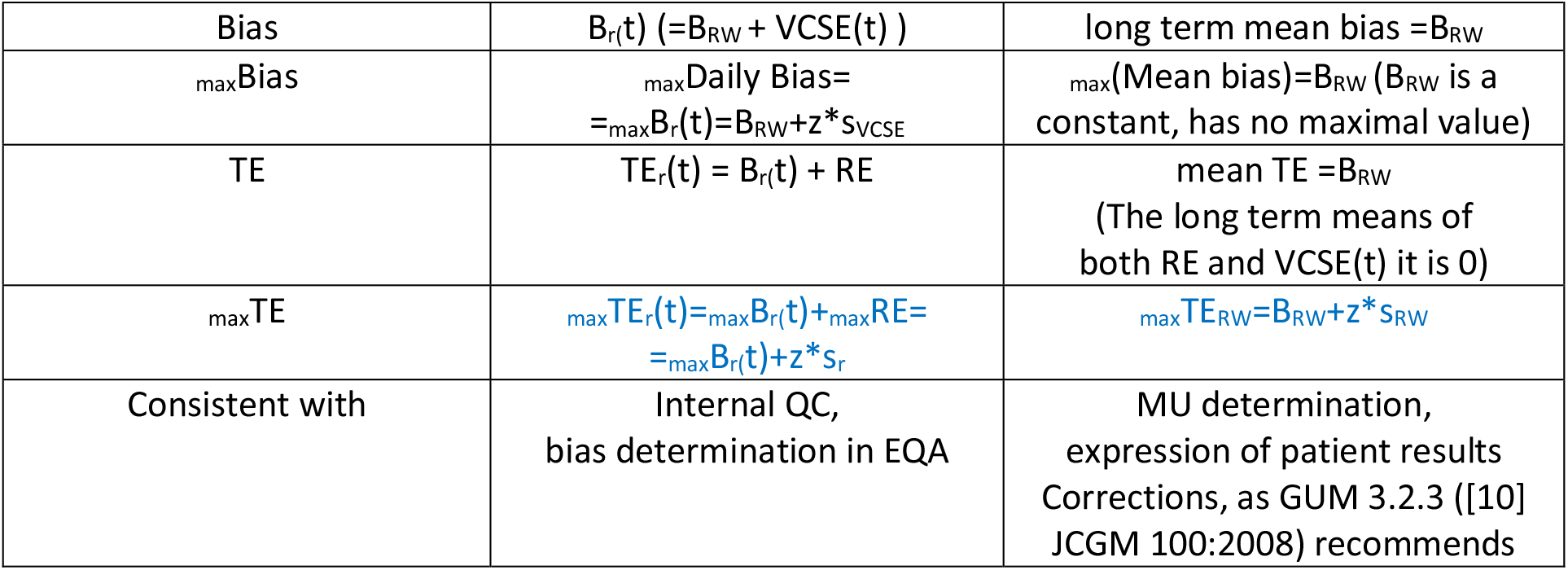
Conditions of determination and area of applicability of the error parameters

Some observations regarding table 1.

- In equations can be used the following pairs: s_r_ and B_r_(t) respectively s_RW_ and B_RW_,
- -Both B_r_(t) and s_RW_ contain VCSE(t) therefore their mix in equations it is equivalent with its redundant use.
- Neither s_r,_ nor B_RW_ contain VCSE(t) therefore their mix is also an error source.
- B_RW_ can be used in repeatability conditions only if it is summed with VCSE(t)
- B_RW_ it is a mean.
- The long-term mean of VCSE(t) and of the RE it is 0. In case of the VCSE(t) the former statement it is true only after several complete cycles.
- The measurement error (TE) in different moments (TE_r_(t)) is different in contrast with the mean measurement error TE_RW,_ which is quasi-constant.
- Although _max_TE_r_(t) and _max_TE_RW_ contain the same three terms, not only their value is different but the two terms also have different meanings. _max_TE_RW_ it is a limit, which is not exceeded with a given confidence by the long term data, while _max_TE_r_(t) it is the same limit applied to the worst day in the same period, in which the bias takes its maximum value.

### What is VCSE(t)?

The VCSE(t) has no definition in VIM. Are systematic error components, therefore by definition are predictable and also variable. While RE changes unpredictable from measurement to measurement, VCSE(t) remains quasi-constant in a given day, influencing all measurement results obtained in that day systematically. But in long-term experiments VCSE(t) becomes a cyclical time-variable function, which repeatedly takes same values after unequal periods. (A period may last even one month).

To understand better the difference between the different bias types, an imaginary, simplified experiment will be presented.

A laboratory has two specialists: A and B. In a given day only one of them is working. Each of them makes volume measurements with his own pipette of 1 ml: P_A_ and P_B_. The two pipettes were metrologically verified. Both had an imprecision expressed as a SD=2μl (CV=0,2%). The inexactity of the two pipettes, expressed as bias it is B_(PA)_=+5 μl (+0,5%) and B_(PB)_=–3 μl (−0,3%). The accuracies of both pipettes are in the allowed limits.

A repeatability experiment in the laboratory (several measurements in the same day) confirmed the metrological results. Specialist A obtained a mean of 1.005 ml, therefore the bias of the day was +0.005ml, and a SD of s_r_=0.002 ml, and 95% of his results were in the 1,005±2*s_r_ interval (between 1.00 1ml and 1.009ml). Next day specialist B obtained a mean of 0.997 ml, and an identical SD of s_r_=0.002 ml, and 95% of his results were in the 0.997±2*s_r_ interval (between 0.993 ml and 1.001 ml). The results of each of them were normally distributed, but they obtained only exceptionally common measurement results. All results obtained in two days in the same laboratory using metrological verified pipettes were not normally distributed, the distribution had two peaks (bimodal), and the results were more dispersed (between 0.993 ml and 1.009 ml). As predictable the SD calculated from all data will be bigger, s_RW_=0.0045 ml, also the mean of results was different: 1.001 ml. The calculated interval for the dispersion of results using s_RW_ in calculation (1.001±2*0.0045) is too large (0.992, 1.010) in comparison with the reality (0.993, 1.009).

The two specialists, surprised by the result repeated the own measurements but they obtained always the same results: the same own mean and the same SD, but the mean calculated from all the results obtained in the laboratory also remain 1.001ml, and the SD=0.0045 ml.

What causes the difference between the SD-s and the means (the biases)? The SD in all days was the same 0.002 ml, the measure of the pure random error, the SD measured in repeatability conditions, s_r_. But in different days the mean of results was different. The mean variation (another variable component) also influenced (increased) the dispersion of results. s_RW_ it is not the SD of the RE but the SD of all variable components. The difference between them is given by the mean (bias) variations.

Each specialist (A and B) measured the bias of the day in repeatability conditions. In each day a different bias value is obtained, therefore the bias measured in repeatability conditions it is a time-variable function B_r_(t). If B_r_(t) it is daily determined and the values are recorded, can be represented the graph of the variations in the past. More than that. If the cause is determined (different pipettes with different biases) and knowing which specialist will work in a future day also can be predicted the value of the bias of that day. In a long period, the measured mean bias (B_RW_ – there are reproducibility within laboratory conditions) will be the average of the daily biases (B_r_(t)). Assuming that the two specialists will work the same number of days, this will be the average of the two possible values (0.997 and 1.005) B_RW_=1.001 ml. The B_r_(t) function will have variations around this value. The difference between the B_r_(t) value, and the mean bias B_RW_ will be the VCSE(t) of the day.

The difference between the two variable error components (RE and VCSE(t)) is the following: while is no way to predict the value of the RE in a future measurement, once determined the daily mean, the B_r_(t) and the VCSE(t) remain quasi-constant in a given day, therefore can be predicted, that in the next measurement will be the same. VCSE(t) it is a systematic error component, and simultaneously a variable one.

The difference between B_r_(t) and the VCSE(t) it is the reference value used to calculate them. While the B_r_(t) is calculated as the difference between the daily mean and the „real” value, the VCSE(t) is calculated relative to the long-term mean of measurements. (The real value of control materials used in internal QC it is unknown. The materials used in internal QC usually do not have certified values, therefore using them only a relative bias (B_r_(t)) value is obtained. Materials used in EQA usually have certified or consent values, accepted as the “real” value, therefore the measured bias values are considered absolute (real) ones.)

s_VCSE_ is a SD calculable from the daily means, the daily bias values or from the VCSE(t) values. The former imaginary experiment gives an explanation also for the variability of the medium term (monthly) means and s_RW_ values calculated from the same data. If only one of the specialists works in a period, the SD measured in “RW conditions” will be approximatively the s_r_ (The mean does not change). This value will be significantly different, if the other specialist also works few days, and more different in comparison with periods in which both work the same number of days. Therefore, using medium term (monthly) s_RW_ and bias cannot be obtained accurate values for TE and MU.

### Causes of the VCSE(t) – shifts in the mean

The former, simplified and imaginary example is a good analogy for what happens in the QC in clinical laboratory, if a very (“perfectly”) stable reagent is used (stable in the period in which the reagent bottle is emptied, and a new bottle is installed). The daily mean remains constant, and the SD (CV) too, therefore the SD will be predictable similar with s_r_ until a (re)calibration is made. A calibration, however has the role to correct errors, it is a measurement, subject of errors, it may introduce variations in the daily means. (If more calibrations are started in short time frames using the same reagents, different calibration parameters are obtained).

One of the causes of the VCSE(t) are the shifts in the daily means, caused by the calibration parameter variations. (E. g. table 2, figure 1). The former statement is sustained also bz the authors of the Nordtest TR 537. (([8] B. Magnusson & al. p. 18). Because the shift is caused by the calibration parameter changes the shift will be similar on both control levels. Its magnitude can be predicted from the old and new calibration parameters (The change of the calibration slope factor F_cal_)

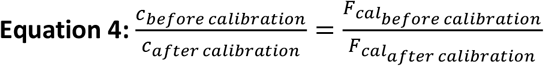

**Table 2:**
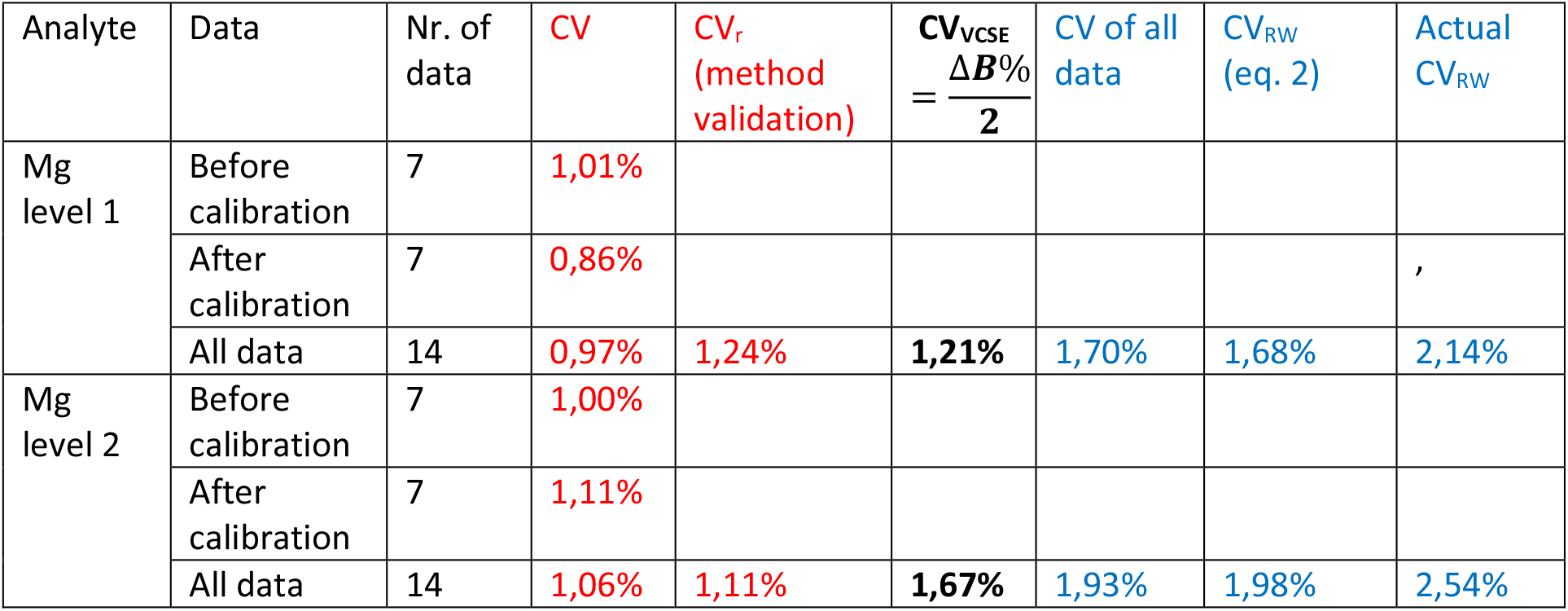
The increase of the s_RW_/CV_RW_ caused by a shift (calibration) can be predicted by equation 2.

**Figure 1:**
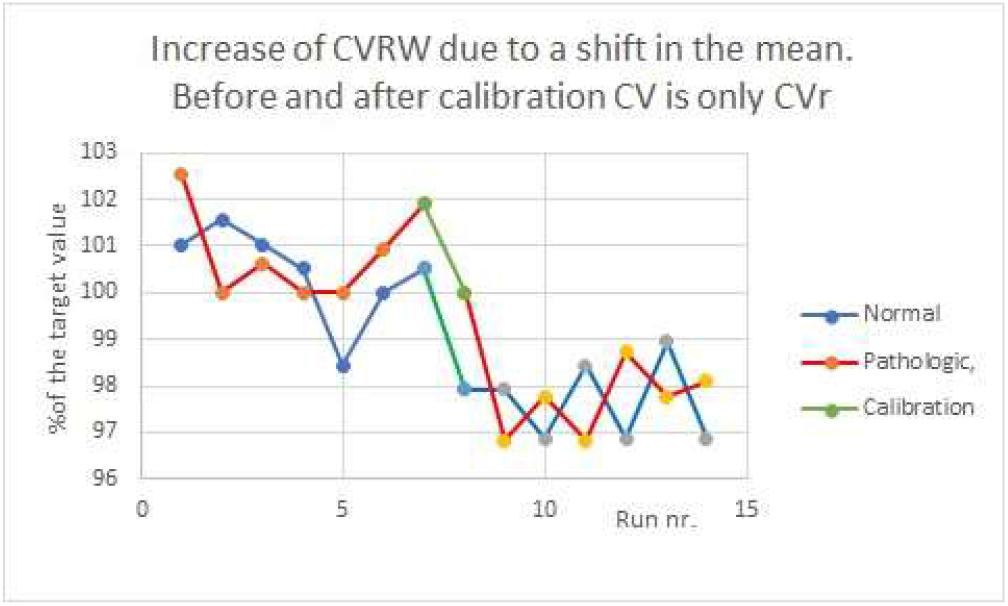
Increase of the CV_RW_ due to a shift in the mean (Mg). CV calculated before and after calibration resemble CV_r._

A typical case (without drift) obtained on a Cobas 6000 analyzer in March 2022 for magnesium it is presented in table 2. For an easier comparison between control levels, the use of CV was preferred instead of SD. The CV calculated before and after calibration (column 4) however were obtained in longer time frames than typical repeatability conditions term data (RW conditions) resemble the CV_r_. CV_VCSE_ was estimated from the mean (relative bias) variation. CV calculated from all data was significantly bigger, resembling CV_RW_ calculated with equation 2. The actual CV_RW_-s (calculated from 168 data) are bigger, than the predicted value for 14 runs because included more reagent changes and calibrations (table 2).

The calculations and predictions are not highly accurate, because of the limits of statistical methods in case of low numbers of data. Even in these conditions, the data sustain the idea that the increase of the CV was caused by the shift in the mean. Figure 1 shows the data on a QC chart.

The presented method to evaluate s_VCSE_ and CV_VCSE_ is only applicable in case of isolated “pure shifts” (with usually known cause), without drifts. The s_VCSE_ obtained can be used only for the given period, and cannot be extended for longer or different time-frames.

In case of more shifts, predictions for the values of the s_RW_ and CV_RW_ need complicated mathematical models. The value of s_VCSE_ can be hardly predicted, the s_VCSE_ must be determined differently. But the former method it is a theoretical proof for its existence, and the influence of the shifts on the dispersion of data.

The influence of the shifts on the SD were confirmed also by computer simulations (2*500 normally distributed data with difference (shift) in the mean).

Shifts in the mean cannot be reduced to calibration parameter changes. Although QC charts are full of shifts-like variations, most of these are only apparent shifts, (without real changes in the daily means), caused by the RE. But also real shifts may happen mostly in case of carry-over (impurification) or „last drops in the bottle” (accelerated degradation) phenomena. The real shifts usually happen simultaneously on both control levels. The role of the internal quality control is to discover them. But even in these cases, the increase of the SD (CV) is caused by the mean variations, the VCSE(t).

### Causes of the VCSE(t) – Drifts in the mean

Bias variations and VCSE(t) cannot be reduced to shifts. Drifts (gradual changes in the daily means caused by reagent property changes-degradation) also contribute to it. Relative short term (few runs) drifts cause quasi-linear changes in the daily mean (also in B_r_(t) and VCSE(t)). Therefore, these variations are predictable from the tendencies.

The equation of the run mean function can be obtained from the parameters of the regression lines (the intercept and the slope) calculated with the method of the least squares from the control data (e. g. using the SLOPE and INTERCEPT functions from Excel). Once determined the equation of the drift, can be determined the value of the mean for each day (run).

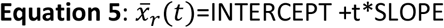

where ‘t’ it is the run number, and 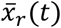 is the daily (run) mean.

The bigger the number of measurements used to calculate the equation, the lower the uncertainty of the estimated values.

From the estimated 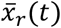 values can be calculated the s_VCSE_. Because the distribution is quasi-uniform s_VCSE_ can also be obtained by dividing the mean (or bias) variation by 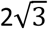. [11] Dividing it by the long term mean value, and multiplying by 100, CV_VCSE_ is obtained for the period of the drift.

The RMS – root mean square – it is the root of the arithmetic mean of squares. The RMS can be calculated from the differences of the measurement results x from the mean of measurements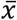(RMS of the residuals= RMS_residuals_)

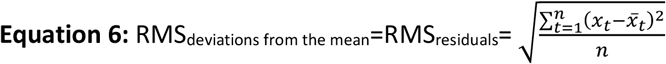

where x_t_ it is the result nr t, 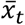 the mean of measurements in the run t, and ‘n’ the numbers of data.

Applying the Bessel’s correction (‘n’, the number of data being substituted with the grades of freedom ‘n-1’[11],) the equation of the root mean square error RMSE is obtained. ([12])

If all the residuals are calculated relative to the long-term mean of all measurements 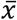, RMSE will be equivalent with a SD, more exactly the equation of the s_RW_ is obtained.

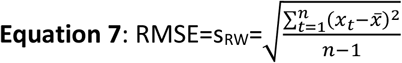

On the graph of any drift can be observed, that the deviations from the daily (run) means 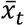 are significantly less than expected from the s_RW_ (CV_RW_) values. Any would be the number of measurements in a given run (day) the dispersion of data in that day (run) will have less SD than s_RW_., because the dispersion of data in long-term RW conditions have two differ sources: the RE and the daily (run) mean variation. Therefore, the RMSE calculated not relative to the long-term mean 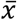, but relative to the daily (run) mean 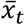 the calculated RMSE will be equivalent with the mean s_r,_ the SD of the pure RE:

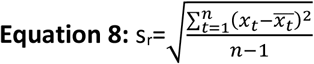

Using the control values expressed as percent of the long-term mean the CV_r_ is obtained.

The obtained value for CV_r_ can be compared with the previously CV_r_ (e. g. measured in the method validation). Using the estimated s_VCSE_ can be calculated the probable s_RW_ for the period of the drift and the actual data for CV_RW_.

A case of 15 run drift (total bilirubin) was chosen for exemplification. Long-term mean values: level 1: 0.88 mg/dl, level 2: 2.92 mg/dl. For easier comparison between levels, the use of CV was preferred (table 3).

**Table 3:** The increase of the CV in reproducibility conditions as consequence of drifts. The measure of dispersion of data around the variable mean (drift) is CV_r_. The increase of the CV of all data is caused by the VCSE(t)

| Measurand | Slope<br>(mg/dl/day)<br>(%/day) | Intercept<br>mg/dl | CV<sub>r</sub><br>(eq. 8) | CV<sub>r</sub><br>(method<br>validation) | CV<sub>VCSE</sub><br>(from<br>$\bar{x}_t$) | CV<sub>RW</sub><br>(All<br>data) | CV<sub>RW</sub><br>(eq. 3) | CV<sub>RW</sub><br>(actual) |
| --- | --- | --- | --- | --- | --- | --- | --- | --- |
| Total<br>Bilirubin<br>Level 1 | 0.0062<br>(0.7%/day) | 0,9046<br>(102.7%) | 1,79% | 1,52% | 3,17% | 3,64% | 3,55% | 3,37% |
| Total<br>Bilirubin<br>Level 2 | 0.0175<br>(0.6%/day) | 2,96<br>(102.8%) | 2,42% | 2,10% | 2,69% | 3,61% | 3,56% | 3,99% |

Using Cochran’s F-test neither between the CV_r_ calculated with equation 5 and the previously determined value obtained in the method validation (table 3, column 4 and 5), nor between the predicted, calculated and actual CV_RW_ values (table 3, last 3 columns) were observed significant differences, (which were caused by the limits of the statistical methods in case of few data) while the CV_r_ and CV_RW_ values were significantly different. Could be concluded, that drifts (mean variations) contribute to the increase of the SD/CV in RW conditions. The graph is presented in figure 2.

**Figure 2.**
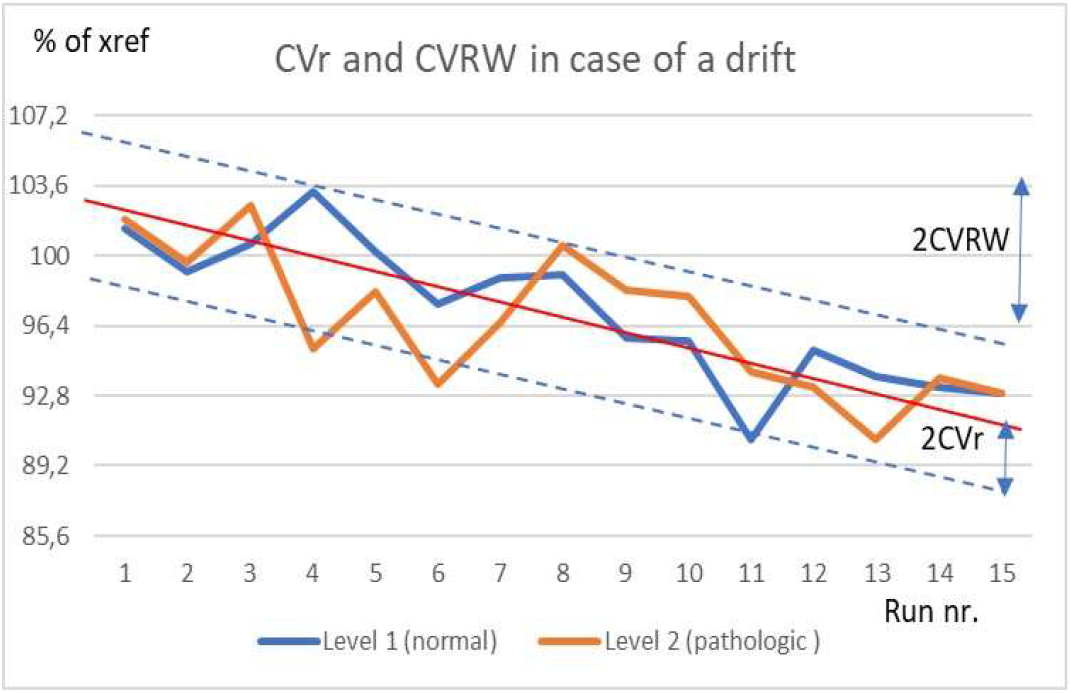
CV_r_ and CV_RW_ in a drift.

Former conclusions were also confirmed by computer simulations. To 1000 normally distributed values a linearly variable bias was added, simulating the drift.

Without doubts, the variations in the daily (run) mean (the VCSE(t)) contribute to the increase of s_RW_ (CV_RW_) in comparison with s_r_ (CV_r_). But in case of several drifts and shifts a mathematical model to calculate the contribution of the VCSE(t) to the dispersion of results cannot be established. In real life cases (the increase of) VCSE(t) and s_VCSE_ must be evaluated from internal QC data. This is the challenge.

While (real) shifts in the mean may have more causes, real drifts are in most cases caused by reagent property changes. (False drifts may be caused by control material degradation). Drifts follow recognizable patterns, therefore are predictable. (The slope may increase in time).

### Real life complex cases: several shifts and drifts

Even in complex cases with several shifts and drifts VCSE(t) can be determined for each day/run, but such determinations have high cost/effectiveness ratio. It is not usual in daily practice, it is justified only for scientific reasons. A single measurement to determine the daily (run) mean has ±z*s_r_ uncertainty. (A single measurement means always repeatability conditions. To determine the uncertainty of the bias the uncertainty of the reference material must be added). Using more measurements in repeatability conditions, the uncertainty of the mean decreases 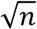 times, where n it is the number of replications. The mean and the bias of that day (B_r_(t)) can be determined with acceptable accuracy.

Repeating this experiment in each day/run in a longer period, the B_r_(t) function can be determined, then s_VCSE_ for that period can be calculated, any would be the number of the shifts and drifts and their cause. An important observation: the VCSE(t) is a cyclical function, the lengths of the cycles depend on the reagent changes, reagent stability and frequency of calibrations, and a cycle may last even more than a month. Also after each calibration the shape of the cycle is different, (because of the different calibration parameters). Therefore, s_VCSE_ determined in relative short time-frames cannot be used for long term decisions.

The CLSI EP15-A3 recommendations for user verification of precision and estimation of bias of the Clinical and Laboratory Standards Institute recommends 5*5 measurements (five runs in each of five days) and an ANOVA single factor analysis for variances. Calculations can be made in EXCEL. (also other software are available). As results three variances are 2obtained. Respecting the notations of the CLSI EP15-A3 V_w_ (variance within runs) is the square of the mean s_r_ (the RMS of the run s_r_-s), V_wl_ (total within laboratory variance), is the square of s_RW_ (s_wl_ in CLSI EP15-A3 notations). In the third one, the V_b_ (variance between runs) can be identified the square of the VCSE, because, according to CLSI EP15-A3, p 23, equation 5, and equation 2 in present study:

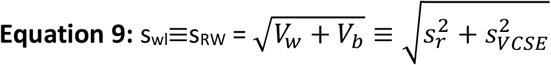

Must be underlined, that s_RW_ (s_wl_) respective s_VCSE_ calculated with EP15-A3 method are only valid for the short time-frame in which they were determined, and cannot be extended for different or longer time frames. The s_wl_ determined in EP15-A3 method significantly under-evaluates monthly quarterly or yearly s_RW_, (and as consequence the s_VCSE_) because each shift, calibration or longer drift will contribute to the increase of the s_VCSE_ and implicit of the s_RW._ While in a five runs (5*5 measurements) experiment (as CLSI EP15-3A recommends) in case of very stable reagents the calculated V_b_ and the corresponding s_VCSE_ can be 0, and implicite s_r_=s_wl_ (≡s_RW_), in case of monthly data never happen. Therefore to exemplify complex cases an unstable reagent (alkaline phosphatase), and to increase the accuracy longer time frame and more measurements in a run was chosen (10*10=100 measurements in 15 days) including also two calibrations. In the days in which a calibration was made, measurements were made two times: once before and also after calibration. Because the bias was not significant in the experiment, the high-level control (target value 246 U/l) was used for measurements. (Because the control material had not certified values, the obtained bias is only a relative bias, not an absolute value.

Experimental data are presented in table 4 together with the calculated parameters: relative B_r_(t) of each run calculated relative to the producer’s target value, each run s_r_, B_RW_ for this period, the mean s_r_, s_VCSE_ and the s_RW_.

**Table 4:** Experimental data to determine VCSE(t) and s_VCSE_ for ALP. Results are given as deviations from the target in U/l.

| Run nr. | 1 | 2 | 3 | 4 | 5 | 6 | 7 | 8 | 9 | 10 |
| --- | --- | --- | --- | --- | --- | --- | --- | --- | --- | --- |
| Day nr. | 1 | 1* | 3 | 5 | 7 | 9 | 9* | 11 | 13 | 15 |
|  | -3.0 | 1.2 | 2.1 | -2.4 | -3.0 | -3.9 | -0.9 | -3.6 | -6 | -8.7 |
|  | -6.9 | 2.7 | -0.3 | -3.9 | -5.7 | -9.9 | 0.6 | -4.5 | -9.9 | -11.1 |
|  | -7.5 | 4.2 | 1.2 | -2.4 | -4.2 | -8.4 | 7.5 | -3 | -8.4 | -9.6 |
|  | -10.2 | 6.6 | -5.4 | 2.1 | 0.6 | -4.5 | -3.9 | -0.3 | -4.2 | -9.9 |
|  | -15 | 6 | 5.1 | 1.2 | -2.1 | -9.3 | 2.1 | 0.3 | -2.1 | -8.4 |
|  | -6.3 | 5.1 | 4.2 | -2.4 | -2.7 | -9.6 | 3.6 | 6.3 | -4.5 | -9.3 |
|  | -11.4 | 5.7 | 3.3 | -1.8 | -1.8 | -3.3 | 0.3 | -4.2 | -13.2 | -14.4 |
|  | -9.3 | 2.7 | 9.6 | 0.0 | 5.4 | -6.3 | 5.7 | 0.9 | -3.3 | -5.1 |
|  | -6.6 | 9.3 | 6.3 | 2.7 | 0.9 | -3.3 | 2.1 | 2.1 | -3.3 | -4.5 |
|  | -4.2 | 10.5 | 3.9 | -2.4 | -3 | -7.2 | 0.0 | 1.8 | -6.6 | -2.7 |
| $B_r(t)$ | -8.04 | 5.4 | 3.0 | -0.93 | -1.56 | -6.57 | 1.71 | -0.42 | -6.15 | -8.37 |
| $VCSE(t)$ | -5,85 | 7,59 | 5,19 | 1,26 | 0,63 | -4,38 | 3,90 | 1,77 | -3,96 | -6,18 |
| run $s_r$ | 3.54 | 2.92 | 4.04 | 2.26 | 3.14 | 2.67 | 3.29 | 3.44 | 3.47 | 3.44 |
| mean $s_r$ = 3.26 | $s_{VCSE}$ = 4.74 | | $s_{VCSE}/s_r$ = 1,45 | | $B_{RW}$ = -2.19 | | $s_{RW}$ = 5.75 | | | |

The results were represented on a classical Stewart-Levey-Jennings graph, designed with SD=6 U/l. The ANOVA (single factor) analysis showed no significant differences between run s_r_ values with 90% confidence. The run s_r_ variations can be explained by statistical uncertainties therefore with 90% confidence s_r_ can be considered time-independent.

On the other hand, a Cochran’s F-test had shown significant differences between all adjacent run means, sustaining, that the increase of the SD in RW versus repeatability conditions is caused by the daily mean variation, the VCSE(t).

The classical Stewart-Levey-Jennings graph it is presented in figure 3. Continuous red lines represent drifts in the mean, dotted red lines shifts caused by calibrations, red points the daily means, blue points the individual results (10 in each run), dotted blue lines the confidence interval of the daily results (±2s_r_), horizontal lines the n*s_RW_ limits.

**Figure 3:**
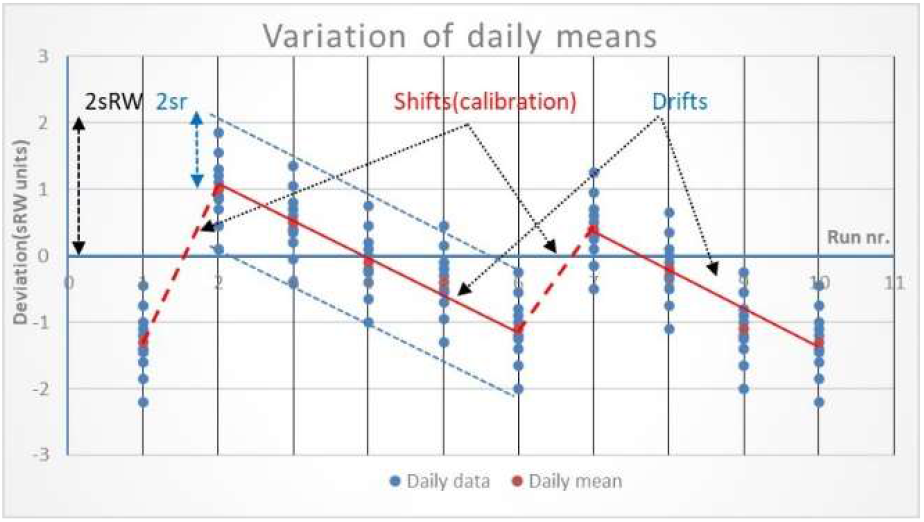
VCSE: Shifts and drifts in the mean (ALP level 2)

On the graph can be observed, that both corrective calibrations caused shifts in the daily means, while between calibrations are drifts caused by reagent property change in time (degradation). While usual control charts have unpredictable variations, apparent shifts in both directions, the daily means have predictable quasilinear variations, sustaining that VCSE(t) is a systematic phenomenon.

### Accurate determination of s_VCSE_

While the determination of s_VCSE_ with the former method is always possible, needs too many measurements, has high cost/effectiveness ratio. A more efficient method is suggested by equation 3. But there are necessary accurate values for s_RW_ and s_r_.

For calculating s_RW_ the internal QC data are available. According to Chi-square distribution ([13], NIST) 3*30=90 data/month would guarantee ±10% imprecision, but only in case of normally distributed data. Because the influence of the VCSE(t) the condition is not respected. Monthly s_RW_ values and also the monthly means have much bigger variations, than expected. [14] B.V. Kumar & T. Mohan, 2018) s_RW_ measured in a given month may be even double of the value determined in another. Only very long term (yearly) control data including several calibrations and reagent (lot) changes are quasi-normally distributed. The computers of the analyzers do not store one year control values. According to the experience and calculations of the author of present study the mean s_RW_ calculated as RMS monthly s_RW_ underestimate the real yearly s_RW_ (calculated from all data in a year). To obtain a correct value, must be added the SD calculated from the monthly means (s_means_):

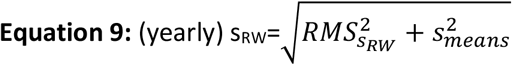

Equation 9 is an analogue of equation 3. Equation 9 was verified by computer simulation. To 10 sets of 100 normally distributed data with variable SD-s variable biases were added, the s_RW_ calculated from all 1000 data was compared with *RMS*_*SD*_ calculated from the SD of each set, and the SD calculated with equation 9. The simulation confirmed the validity of equation 9.

Usually s_r_ is calculated from a replication experiment, and extra measurements must be made. According to the Chi-square distribution not even 25 replications guarantee less than 25-30% imprecision. But the time-independent character of s_r_ permits its determination also from long term data.

A SD is calculated from the deviations from the mean with equation 7. A SD of a difference of values with the same SD it is 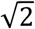 times bigger, than the SD of the individual values. Calculating the SD of differences obtained in repeatability conditions, then dividing the result with 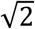 is obtained the value of s_r_. Usually two control measurements are made in the same run, but using different control materials, (more control levels) which in absolute values are incomparable because their different values. But the control results expressed as percent of their mean value do not depend any more of the concentration (activity) of the control material.

Monthly CV_RW_ and s_RW_ values have significant variations. For the same reason, CV_RW_ values determined in the same month may be significantly different. If long-term control data are not normally distributed, control values obtained in repeatability condition are (they are not influenced by the VCSE(t) function). Also s_r_ is proportional with concentration, therefore CV_r_ values are relative constant in the measurement range (excepting the values around to the limit of quantitation). ([4] A. B. Vandra, 2014, p. 144) A similar linear dependency of the measurement uncertainty was described in Eurachem/CITAC Guide ([15] QUAM, 2012, p. 117, and figure E.5.1., p. 120).

Expressing the control values as percent of their mean values, calculating the SD of the differences between different control levels, then dividing the result by 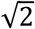 a mean CV_r_ is obtained.

Multiplying CV_r_ with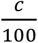, where c it is the concentration of the control material, can be obtained the s_r_ values for both controls. Table 5 presents an example of such a calculation in case of urea:

**Table 5:**
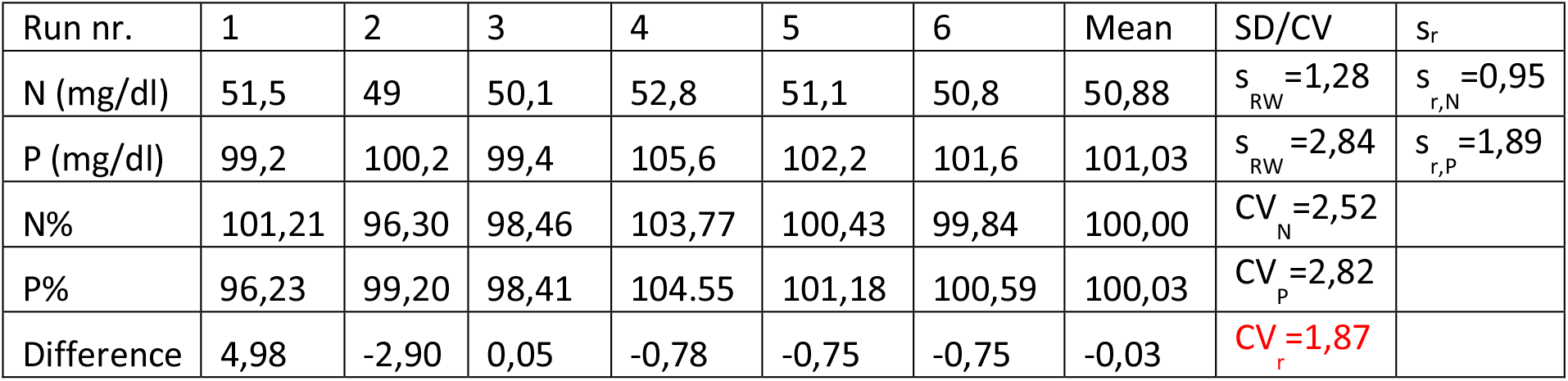
Example of calculation of CV_r_ and s_r_ from long term data (6 runs) for urea. 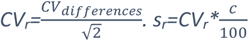

(A similar method to determine s_r_ from long-term obtained data pairs is described in Nordtest TR 537, first edition, appendix 5 and 6, page 38-39, but instead of 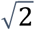 they divided CV_differences_ with 1,128 as recommended in appendix 8, page 41 ([16] B. Magnusson & al. 2003). In the 4^th^ 2017 edition appendix 5 and 6, page 41-42, this error was corrected, and the correct division by 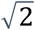 is used, however is not specified ([8] B. Magnusson, 2017).

The former method to determine CV_r_ from long term control data was simulated on computer. RE was simulated by 1000 normally distributed data, VCSE(t) by a sinusoidal function. The results confirmed the theoretically deduced 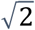 division factor, against the erroneously recommended 1,128.

The CV_r_ obtained with this method contains additionally the influence of the value variations of the control material (reconstitution, depositing, homogenization, etc. errors). The former method permits to calculate s_r_ from hundreds of data pairs, therefore is more accurate than the value obtained from replication. Substituting s_r_ and s_RW_ obtained in the former conditions in equation 3 an acceptable accurate value for s_VCSE_ can be obtained.

### Differences between the three error components: CCSE (BRW), VCSE(t), and RE

The comparison between the three error components it is presented in table 6:

**Table 6:**
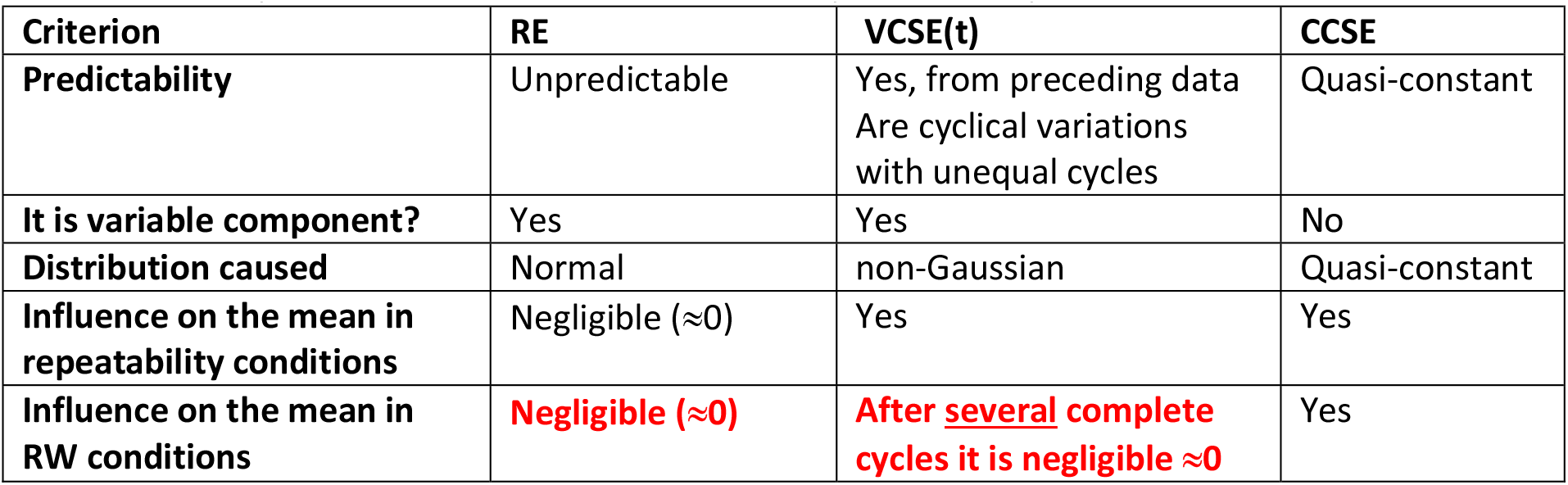
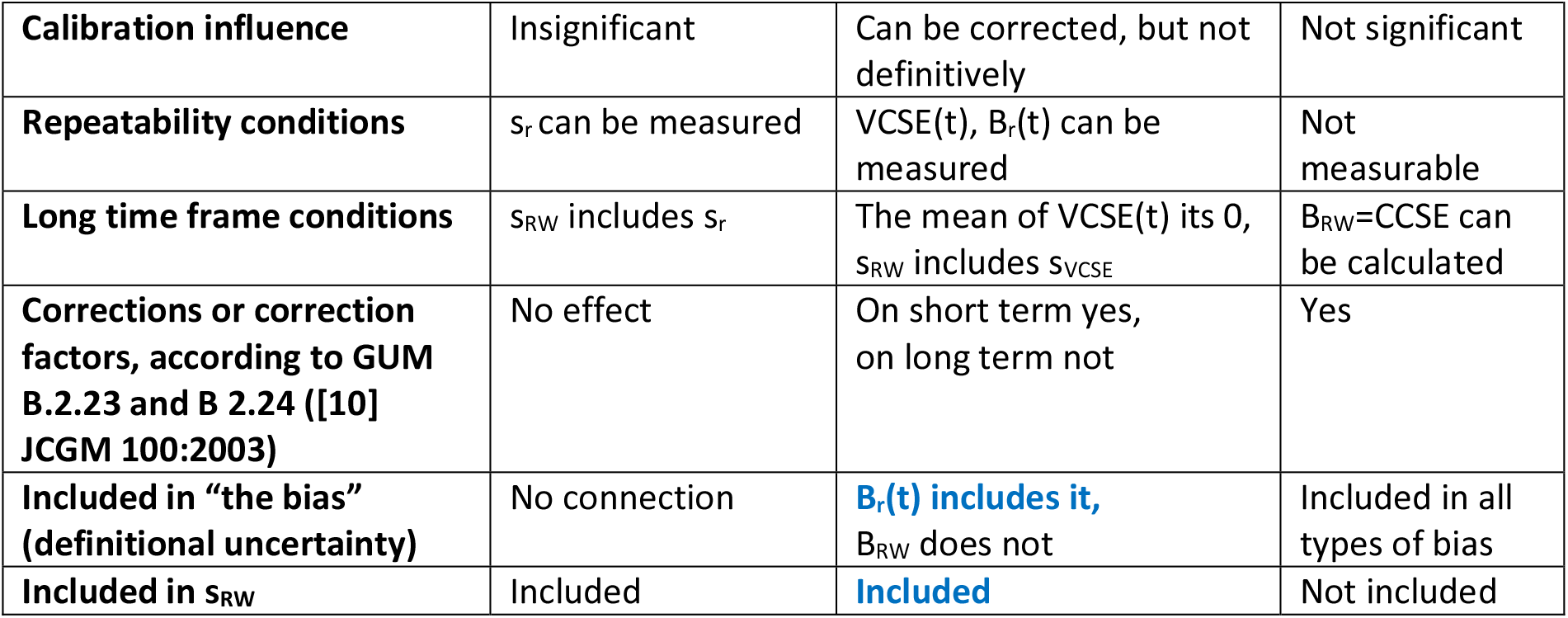
Differences and similarities between the error components. The main source of confusion is highlited in red, the risk of redundant use in blue.

There are too many differences between the VCSE(t) and the other error components (RE, CCSE) to ignore them. It is not erroneous practice, to measure two error components together, and to include VCSE(t) in other components (s_RW_, B_r_(t)) if, and only if, we are conscious that these error parameters contain them. But for respecting this condition, must be known the origins of the equations, which contain the error components separated, and also the risk of their redundant use.

### Can be VCSE(t) transformed into random phenomena?

According to E. Theodorsson & al [5]).

“*Short term systematic effects in time are transformed into random effects*” ([2], E Theodorsson, & al. 2014)

The former erroneous statement is based on a confusion between random and variable components. “Variable systematic” is not equivalent with “random.” In short term the difference is obvious. In longer time frames is more complicated. Because the relative long cycles of variation of the VCSE(t) are unequal, predictions for the distant future cannot be made. But if the daily means are determined and recorded, can be predicted, that a patient sample in the distant past was influenced by the same systematic error, as the controls of that day. But neither the recorded daily means help us to predict the RE of a patient sample in the past. Knowing the tendencies, (drift) also for near future can be made predictions for the bias of next day. If the reagent is stable, until a new calibration (or reagent change) is made the bias remains predictable. The impossibility to make predictions for distant future it is a necessary, but not sufficient condition to consider VCSE(t) transformed into a random phenomenon.

Something transformed neither exist anymore in its original form, nor is measurable, nor its properties can be observed. The melted ice loose its form, and the frozen water cannot be measured by volumetric methods (e.g with a pipette). But the properties of the VCSE(t) influence the distribution of long-term data, which are also not normally distributed (as also VCSE(t) is not). The other criterion it is the measurability. But s_VCSE_ has no sense in short term, in repeatability condition, only in RW condition can be determined.

Therefore, it can be concluded, that no transformation happens, only the influence of the RE and VCSE(t) on the distribution of data are measured together, when s_RW_ is determined.

When calculating MU, it is not a big deal to make difference between “transformed” and “measured together”. Anyway, RE and VCSE(t) are summed, and included in the total uncertainty. But there is a condition, which must be strictly respected: concomitantly VCSE(t) must be eliminated from the systematic component (to use the uncertainty of the mean bias in calculations instead the vaguely defined “bias uncertainty”).

But in case of internal quality control the difference is crucial. The role of the internal QC it is to detect risky increases of the VCSE(t). In a hidden form (included in B_r_(t) or s_RW_) it is a game of blinds.

## Conclusions

The actual paradigm of the QC in clinical laboratory must be revised. In first step must be made a clear difference between the bias measured in repeatability respective in RW conditions, as we are accustomed in case of SD-s. Also, the same principles for notation must be used for them (B_r_(t), B_RW_). However, VIM does not define systematic error subcomponents, to be consistent with the definition of VIM, SE must be separated into a constant (CCSE) and a variable subcomponent (VCSE(t)). The VCSE(t) it is the error subcomponent, which links the error parameters (Bias, SD) measured in repeatability, respective RW conditions. The bias of the moment (B_r_(t)) and s_RW_ contain VCSE(t) in hidden form.

It is important to separate the VCSE(t) from other error component is equations to avoid its redundant use. TE and MU equations must be reevaluated from this point of view, because both “the bias” and RMS_Bias_ may contain definitional uncertainties, including VCSE(t) in hidden form, which is present also in s_RW_. (The deep analyze of these redundancies is not the task of the present study).

The error parameters determined in a given condition (repeatability or RW) must be consistent with the conditions of the predictions they are used. To calculate MU the B_RW_ and s_RW_ must be used, in internal QC decisions s_r_ and B_r_(t). The last conclusion it is in contradiction with the actual recommendation, which must be radically revised. (The actual used Westgard rules cannot be used with s_r,_ a new rule system is necessary).

To avoid the uncertainty in the definition of “the bias” (which one?) VCSE(t) must be defined in VIM, and highlighted in equations, to become conscious which error parameters contain it, even if later they are measured together or are included in other error parameters (e. g. s_RW_, B_r_(t)). The VCSE(t) have many properties which makes them distinct from RE and CCSE, therefore deserves a separate notation, and place in equations. One of these properties it is the not Gaussian distribution of the daily means.

The VCSE(t) consist of shifts and drifts in the daily mean, and the role of the QC is to detect them. Highlighting them in equations, makes the QC decisions more conscious.

For the accurate determination of the SD-s (s_r_, s_RW_ and s_VCSE_) must be used yearly control data, even s_r_ can be determined from long term data with the method described.

There is a need for a new, more accurate internal quality control system, based on CV_r_, instead s_RW_, because internal quality control decisions are taken in repeatability conditions, therefore the error parameters which are consistent with these decisions must also be determined in repeatability. This new quality control system must have a fundamentally different rule system, based on different principles and postulates. The actual used Westgard rule system fails when using Levey-Jennings charts designed with s_r_ or CV_r_.

## Data Availability

All data produced in the present work are contained in the manuscript.

## Abbreviations

VIM: International Vocabulary of Metrology
QC: Quality Control
EQA: External Quality Assessment
MU: measurement uncertainty
TE: total measurement error
RE: Random error component
SE: Systematic error component
VCSE(t): Variable Component of Systematic Error in the moment t (a time-variable function)
CCSE: Constant Component of Systematic Error
SD: Standard Deviation (in general)
s_r_: SD measured in constant, repeatability conditions
s_RW_: SD measured in variable, reproducibility within laboratory conditions
s_VCSE_: the SD calculable from the daily(run) mean (bias, VCSE(t)) values
s_mean_: the SD calculable from the monthly means
CV: Coefficient of Variation, the SD expressed as percent of the mean of measurements
CV_r_: CV measured in constant, repeatability conditions
CV_RW_: CV measured in variable, reproducibility within laboratory conditions
CV_VCSE_: CV of the VCSE(t), s_VCSE_ expressed as percent of the target value
B: bias
B%: bias expressed as percent of the long-term mean of measurements
∆ *B*%: percent expressed bias variation in a shift (percent of the long-term mean value)
B_r_(t): (short term or within day) bias measured in repeatability conditions in the moment t (a time-variable function)
B_RW_: long term mean bias, measured in RW conditions, a constant
RMS: root mean square - the root of the arithmetic mean of squares (not to be confused with the RMSE)
_max_□ (max index before): the maximum value of a parameter in given conditions
z: coefficient of confidence
c: concentration or activity of a measurand
n: number of measurements
t: time or moment, expressed as run number
F_cal_: the slope factor of the linear calibration curve
x: (control) measurement result
x_t_: (control) measurement result in the moment t
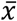: mean of measurements
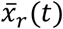: the daily (run) mean in the moment t (repeatability conditions)
CLSI: Clinical and Laboratory Standards Institute
ANOVA: analysis of variance
V: variance, the square of the SD
V_w_: Variance within run (notation in CLSI EP15-A3), the square of s_r_
V_b_: Variance between runs (notation in CLSI EP15-A3), the square of s_VCSE_
V_wl_: Variance within laboratory (notation in CLSI EP15-A3), the square of s_RW_

## Disclaimer

To the present no other persons contributed except the author

The present study was neither funded nor funded

No patient data was used in the present study

## References

[1] Rouvim Kadis: Evaluation of measurement uncertainty in analytical chemistry: related concepts and some point of misinterpretation. Dissertationes chimicae univeritates Tartuensis, Sept. 2008. https://www.researchgate.net/publication/277054223_Evaluation_of_measurement_uncertainty_in_analytical_chemistry_related_concepts_and_some_points_of_misinterpretation

[2] Elvar Theodorsson, Bertil Magnusson, Ivo Leito: Bias in Clinical Chemistry, Bioanalisys, vol 6, no. 21, published online 8 dec 2014, https://www.future-science.com/doi/10.4155/bio.14.249

[3] Churchill Eisenhart: Realistic Evaluation of the Precision and Accuracy of Instrument Calibration Systems. Journal of Research of the National Bureau of Standards-C, Engineering and Instrumentation 67C(2): 161–187, April-June 1963.

[4] Atilla B. Vandra: Incertitudini… în lumea incertitudinii. Deplasarea. (Uncertainties… in the World of Uncertainty. The Bias). Revista Română de Laborator Medical, 35, September 2014, p. 143

[5] Wytze P. Oosterhuis, Hassan Bayat, David Armbruster, Abdurrahman Coskun, Kathleen P. Freeman, Anders Kallner, David Koch, Finlay Mackenzie, Gabriel Migliarino, Matthias Orth, Sverre Sandberg, Marit S. Sylte, Sten Westgard and Elvar Theodorsson: “The use of error and uncertainty methods in the medical laboratory” - The concepts of bias and imprecision, Clinical Chemistry and Laboratory Medicine (CCLM), vol. 56, no. 2, 2018, pp. 209–219. https://doi.org/10.1515/cclm-2017-0341,https://www.degruyter.com/document/doi/10.1515/cclm-2017-0341/html#j_cclm-2017-0341_ref_030_w2aab3b7c59b1b6b1ab2b3c30Aa

[6] Ivo Leito, University of Tartu: LC-MS method validation: Bias and its constituents. https://sisu.ut.ee/lcms_method_validation/51-Bias-and-its-constituents

[7] VIM: Definitions with informative annotations. https://jcgm.bipm.org/vim/en/index.html

[8] Bertil Magnusson, Teemu Näykki, Håvard Hovind, Mikael Krysell, Eskil Sahlin: Handbook for calculation of measurement uncertainty in environmental laboratories (NT TR 537 – Edition 4), page 10, http://www.nordtest.info/wp/2017/11/29/handbook-for-calculation-of-measurement-uncertainty-in-environmental-laboratories-nt-tr-537-edition-4/

[9] Clinical and Laboratory Standards Institute: (R. Neill Carey & al.): User Verification of Precision and Estimation of Bias; Approved Guideline—Third Edition, EP15-A3, 2014, vol. 34 nr. 12, last correction, November 2022, https://clsi.org/standards/products/method-evaluation/documents/ep15/

[10] JCGM 100:2008. Evaluation of measurement data – Guide to the expression of uncertainty in measurement https://www.bipm.org/documents/20126/2071204/JCGM_100_2008_E.pdf/cb0ef43f-baa5-11cf-3f85-4dcd86f77bd6

[11] https://en.wikipedia.org/wiki/Standard_deviation

[12] GraphPad: Standard deviation of the residuals, https://www.graphpad.com/guides/prism/latest/curve-fitting/reg_standard_deviation_of_the_resi.htm

[13] Clinical and Laboratory Standards Institute. User verification of precision and estimation of bias; approved guideline-3rd (ed) Wayne, PA, USA: CLSI; 2014. CLSI document EP15-A3. https://clsi.org/media/3398/ep15a3e_sample.pdf

[14] NIST Sematech: Engineering Statistics Handbook.1.3.5.8 Chi-Square Test for the Standard Deviation http://atomic.phys.uni-sofia.bg/local/nist-e-handbook/e-handbook/eda/section3/eda358.htm

[15] B. V. Kumar, T. Mohan: Sigma metrics as a tool for evaluating the performance of internal quality control in a clinical chemistry laboratory, Tables 3 and 4, J Lab Physicians. 2018 Apr–Jun; 10(2): 194–199., https://www.ncbi.nlm.nih.gov/pmc/articles/PMC5896188/

[16] S. L. R. Ellison and A. Williams (Eds). Eurachem/CITAC guide: Quantifying Uncertainty in Analytical Measurement, (3rd edition, 2012) p 120, Fig E 5.1, Available from https://www.eurachem.org. https://www.eurachem.org/images/stories/Guides/pdf/QUAM2012_P1.pdf

[17] Bertil Magnusson, Teemu Näykki, Håvard Hovind, Mikael Krysell, Eskil Sahlin: Handbook for calculation of measurement uncertainty in environmental laboratories (NT TR 537) first edition, 2003, Nordtest, Tekniikantie12, FIN-02150Espoo, Finland

